# COVID-19 gender difference pattern in Iranian population compared to the global pattern; a systematic review and meta-analysis

**DOI:** 10.1101/2021.05.23.21257692

**Authors:** Misagh Rajabinejad, Hossein Asgarian-Omran

## Abstract

The coronavirus disease 2019 (COVID-19) pandemic has highlighted Sex-related immune responses. In this review, gender differences in seroprevalence, severity, mortality, and recovery in the Iranian population were systematically compared to the COVID-19 global pattern. This compressive meta-analysis was conducted on studies published up to April 1, 2021, examining seroprevalence in the general population as well as disease outcomes in hospitalized patients. Data were analyzed based on gender to determine differences between men and women in COVID-19. The PubMed, Scopus, Google Scholar, WOS, medRxiv, and bioRxiv were searched. The odds ratio (OR) was calculated based on the random-effects model, with a corresponding 95% confidence interval (CI), according to the number of participants reported in papers. Subgroup analyses were performed according to the age, antibody isotype, and detection assay. Overall, 61 studies with 225799 males and 237017 females were eligible for meta-analysis. Seroprevalence was 1.13 times higher (95% CI: 1.03, 1.24), mortality was 1.45 times higher (95% CI: 1.19, 1.77), and severity was up to 1.37 times higher (95% CI: 1.13, 1.67) in males than those of females in the general population across the globe. Mortality was higher in Iranian patients up to 26% in men (95% CI: 1.20, 1.33), but no significant difference was observed between disease severity and serum prevalence between men and women. Besides, the rate of recovery was 29% (global pattern) and 21% (Iran pattern) lower in males than in females. The results of subgroup analyses for seroprevalence were not significant for the age, antibody isotype, and detection methods. The results of our meta-analyses showed that the patient mortality and recovery patterns are similar in Iran and other countries in the context of gender differences, and the disease is more fatal in men.

## 1. BACKGROUND

It has been almost a year and a half since the first case of the coronavirus disease 2019 (COVID-19) was reported in Wuhan, China. In January 2020 the World Health Organization (WHO) declared it as a Public Health Emergency of International Concern, and shortly thereafter in March 2020, it was officially declared as a pandemic (1). The cause of this pandemic is a virus, with an unclear origin, from the coronavirus strain called severe acute respiratory syndrome coronavirus 2 (SARS-CoV-2) (2). This virus has a single-stranded RNA genome encoding several open reading frames, including structural proteins spike (S), matrix (M), envelope (E), and nucleocapsid (N) (3). Despite the start of vaccination in most parts of the world, many countries, including Iran, are still severely affected by the high prevalence of the disease. The unknown pathogenic and immunological dimensions of SARS-CoV-2 have caused a very high amount of mortality.

One of the most important key points of this pandemic is to understand the individual differences in disease severity and mortality. Although the COVID-19 pandemic has become a global infodemic, many aspects of the disease remain unknown, and even serious disagreements and contradictions have arisen among scientists on some issues. From the beginning, reports showed a higher mortality rate in men than women, highlighting the role of chromosome X in host immune response (4). It was initially reported that the reasons for this discrepancy are stronger adaptive and innate immune responses in women, and even better antiviral responses such as early production of interferons (5, 6); but over time, controversy arose over COVID-19 gender difference (7). Preliminary studies have suggested that estrogen, 17β-estradiol, may play a protective role by regulating ACE2 expression as the major cell entry receptor for SARS-CoV-2. However, the results of studies on the effects of estrogen on ACE2 expression are controversial and cannot be commented on with complete certainty (Table 1). For this reason, researchers are now debating the effects of estrogen on regulation of ACE2 and how it affects the pathogenesis of SARS-CoV-2. Another factor that is somewhat questionable is the effect of interferon responses on virus inhibition and its role in the gender differences in COVID-19. Recent studies show that only early interferon type 1 responses can control the virus pathogenesis (8). In addition, various studies have been performed on the effect of interferons on ACE2 expression, and these results are slightly different (Table 2). However, most of these studies have reported that type 1 and 2 interferons in both *in vivo* and *in vitro* environments may be associated with increased expression of ACE2, whereas how interferon can play a role in gender differences is still controversial.

**Table 1.** Effect of estrogen on ACE2 expression.

| **Year** | **Sample type** | **ACE2 expression** | **Reference** |
| --- | --- | --- | --- |
| **2020** | Differentiated normal human bronchial epithelial cells | Decreased | (27) |
| **2020** | Mouse thymus | Increased | (28) |
| **2020** | Female Wistar rats | Increased | (29) |
| **2020** | Ovariectomized CD1 mice | Decreased | (30) |
| **2020** | Human airway smooth muscle cells | Decreased | (31) |
| **2020** | Human umbilical vein endothelial cells and C57BL/6 mice | Decreased | (32) |
| **2019** | Obese female C57BL/6 mice | Increased | (33) |
| **2017** | Human atrial tissue from male donors | increased | (34) |
| **2015** | Human umbilical vein endothelial cells | Increased | (35) |
| **2015** | HF-fed OVX female mice | Increased | (36) |
| **2014** | Ovariectomized fructose fed female Wistar rats | Increased | (37) |
| **2014** | OVX Sprague-Dawley female rats and Sprague-Dawley male rats | Increased | (38) |
| **2013** | Female OVX-mRen2.Lewis's rat | Decreased | (39) |
| **2012** | OVX HF-fed female C57BL/6 mice | Increased | (40) |
| **2011** | Female OVX Sprague-Dawley rats | Increased | (41) |
| **2009** | DOCA-salt model of hypertension in female OVX Sprague-Dawley rats | Increased | (42) |
| **2008** | Female OVX Sprague-Dawley rats | Increased | (43) |
| **2008** | Female ApoE<sup>-/-</sup> mice, with or without ERα on a C57Bl6/J genetic background | Decreased | (23) |
| **2007** | Pregnant Sprague-Dawley rats | Increased | (44) |

**Table 2.**
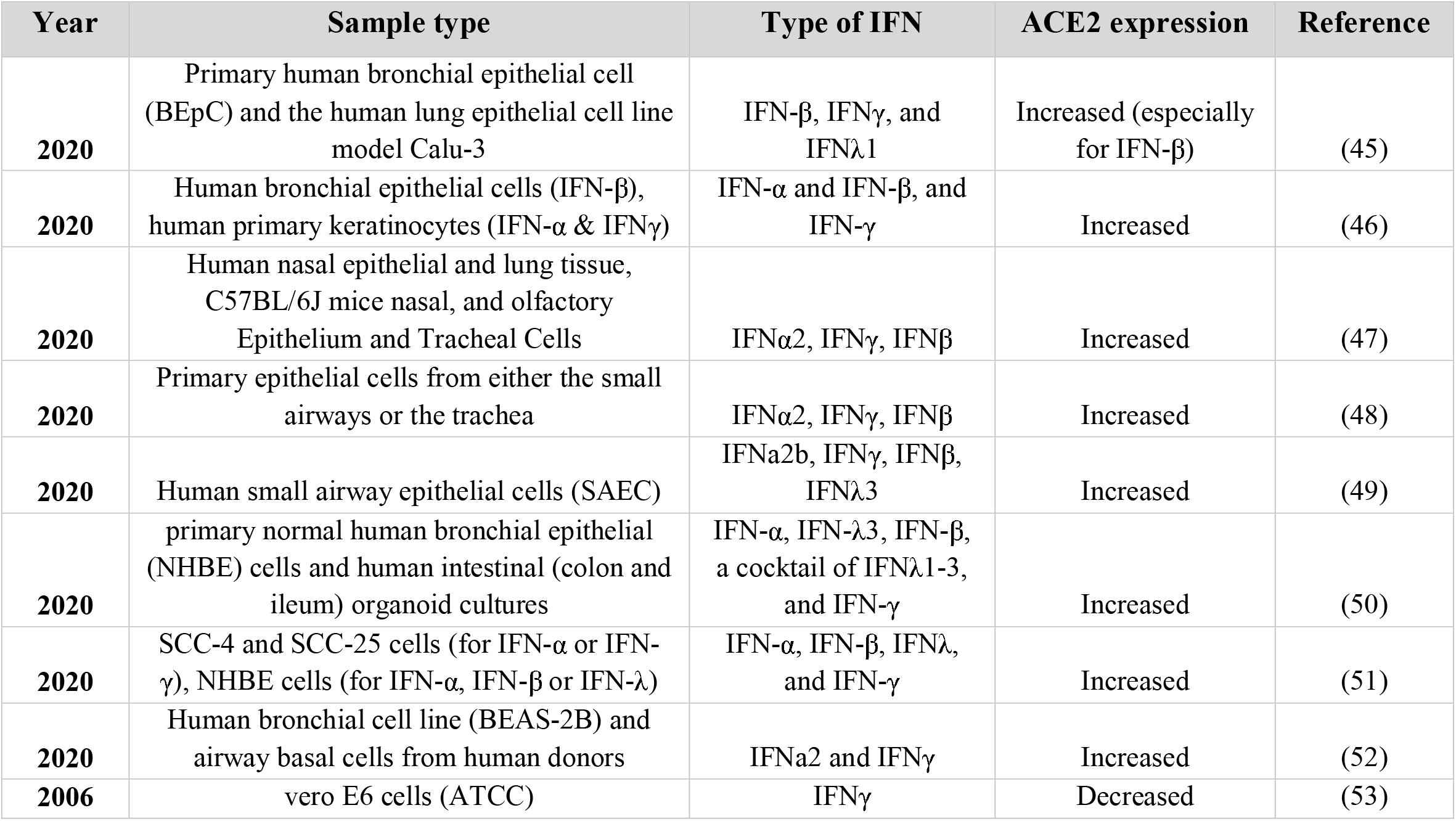
Effect of IFN on ACE2 expression.

COVID-19 can be compared to an iceberg which more than half of it is underwater and invisible (9). Therefore, if the goal is to evaluate the mortality, severity, or pathogenicity of coronavirus, the underwater part should also be clearly seen. Seroprevalence studies can give a good view of the coronavirus infection in the general population and estimate the frequency of challenged people (10). These types of studies identify and report the number of people who have a negative RT-PCR test but a positive serology test for anti-coronavirus IgM or IgG. Summarizing this group of information can provide a very good view of the number of asymptomatic patients in the population, and may eventually be used to refute or confirm the controversial theory of herd immunity (11).

The most important step in treatment and vaccination is to fully understand all differences in the target population, and perhaps the first step in achieving personalized medicine is to know the gender characteristics in treatment and vaccination. The potential gender differences in COVID-19 pathogenicity can also affect the effectiveness and safety of ongoing vaccination. For these reasons, and of course the lack of a comprehensive systematic study to examine gender differences in seroprevalence, mortality, severity, and recovery of COVID-19, the present study was designed and conducted. This study tried to focus on the differences between women and men in the general population and provide information that would help to care, follow-up, and monitoring of high-risk groups. Comparing the results of the global meta-analyses and the results of studies conducted in Iran can provide a very good insight for researchers in the context of herd immunity and gender-related mortality.

## 2. METHODS

### 2.1 Study design

Systematic review and meta-analysis on gender-related COVID-19 outcomes were conducted according to the standard protocols. The review protocol was registered in PROSPERO (CRD42020216637).

### 2.2 Inclusion criteria

#### 2.2.1 Types of studies

All published and/or online papers as of April 1, 2021, that examined gender-based seroprevalence, severity, mortality, or patient recovery in COVID-19 were included in our study. The types of studies considered for inclusion were cross-sectional, case-control, and cohort studies. Papers from any country concerning gender-specific outcomes were included if they were written in English. We only included studies that reported the exact number or percentage of participants by gender (sex-disaggregated), as well as the exact type of detection method and the population studied. For papers reporting both IgG and IgM, only IgG data were included in the total seroprevalence analyses. Studies conducted on healthcare workers, patients of other disorders or infections, as well as those participants with unknown exposure were excluded to assess the true effect of gender on COVID-19 seroprevalence. However, to evaluate other outcomes, including severity, mortality, and recovery of patients, meta-analyses were performed on studies with COVID-19 confirmed cases. Reviews, meta-analyses, case reports, abstracts, and overlapping papers were excluded. Moreover, the references of relevant studies were reviewed to ensure the absence of the missing articles. Besides, due to the significant difference in the rate of vaccination of the population in Iran and some developed countries, as well as the effect of vaccination, especially on seroprevalence and mortality, only papers whose sampling date was until the end of 2020 and before general vaccination were included in global meta-analyses.

#### 2.2.2 Types of participants

Eligible studies for seroprevalence analysis have to include participants of any age or sex from the general population (local population, residents, households, and blood donors). Eligible studies for severity, mortality, and recovery have to include patients with confirmed COVID-19 by molecular diagnostic methods (RT-PCR) and the information of their death or recovery and discharge was clearly reported. We also included studies in which participants did not have any other medical disorders, such as autoimmunity, primary or secondary immunodeficiency, malignancy, allergy, and acute or chronic infections (except COVID-19). We didn’t use any restrictions concerning age, sex, and COVID-19 related comorbidity.

#### 2.2.3 Types of clinical outcomes

1. Gender-related differences in seroprevalence in Iran compared to the global pattern (frequency of individuals who are seropositive and RT-PCR negative for COVID-19 in the general population).
2. Gender-related differences in COVID-19 severity in Iran compared to the global pattern (frequency of COVID-19 male and female patients, whose disease conditions were reported to be severe and critical).
3. Gender-related differences in COVID-19 mortality in Iran compared to the global pattern (frequency of deceased male and female COVID-19 patients).
4. Gender-related differences in recovery and discharge of COVID-19 patients in Iran compared to the global pattern (frequency of recovered or discharged male and female COVID-19 patients).

### 2.3 Electronic searches

PubMed (1^th^ April 2021), Scopus (1^th^ April 2021), Google Scholar (3^rd^ April 2021), Web of Science (WOS-4^th^ April 2021), and two preprints servers (medRxiv and bioRxiv) were searched using the Medical Subject Headings (MeSH) terms “male”, “female”, “men”, “women”, “sex”, “gender”, “corona”, “COVID-19”, “Cov2”, “SARS”, “SARS-COV-2”, “SARS-2”, “SARS-corona”, “severe acute respiratory syndrome”, “mortality”, “morbidity”, “death”, “clinical course”, “clinical presentation”, “intensive care”, “hospital stay”, “seroprevalence”, “seroincidence”, “seroconversion”, “seronegative”, “seropositive”, “seroepidemiologic”, “serologic”, “serosurvey”, “antibody”, “attack rate”, “severity”, “critical”, “recovery”, and “discharge”. The references listed in related publications were also searched. There was not any restriction on searching.

### 2.4 Data collection and analysis

#### 2.4.1 Selection of studies and data extraction

Both two authors (MRN, HAO) independently screened all search records and identified those that were fully published and fulfilled the inclusion criteria. To homogenize the data as much as possible, only studies that reported the exact number of participants in both male and female groups were included.

#### 2.4.2 Measurements of treatment effects

All reported seroprevalence, severity, mortality, and recovery data were pooled to investigate gender-related outcomes. Statistical analysis was performed using the Cochrane software Review Manager v5.3 (RevMan v5.3). For the dichotomous outcome, the odds ratio (OR) was calculated based on the random-effects model, with a corresponding 95% confidence interval (CI), according to the number of participants reported in the papers. For continuous variables, standardized mean difference and *p*-value were calculated according to the mean, standard deviation (SD), and the number of participants reported in the papers. We only performed meta-analyses for the outcomes that were homogenous, and the number of participants was clearly reported by gender. Subgroup analyses were performed for seroprevalence and mortality based on the age group. Type of antibody and detection method-based subgroup analyses were also performed for seroprevalence. The unadjusted (crude) data were used in our analyses to prevent heterogenicity in different studies. The bias risk of the included studies was assessed by the Newcastle-Ottawa Scale (NOS) (12). The statistical heterogeneity between trials was also evaluated using the Q-test of heterogeneity and the I^2^ test of inconsistency. Any disagreements were resolved by discussion among the review authors.

## 3. RESULTS

### 3.1 Search results

The study selection process for this systematic review is shown in Figure 1 as the PRISMA flow chart. After the initial searches, 8532 records (PubMed 1668, Scopus 1590, Google Scholar 2746, WOS 1039, MedRxiv, and BioRxiv 1489) were identified. After removing duplicates, reviews, and unrelated papers, 61 papers that matched our inclusion criteria were found. In 20 out of these 61 studies, seroprevalence was reported in the general population (18 global, 2 Iran). The remaining 41 papers reported the severity, mortality, and recovery by gender. Of these 49 articles, 19 (16 global, 3 Iran) were included for severity analysis, 27 (16 global, 11 Iran) for mortality analysis, and 19 (8 global, 11 Iran) for recovery analysis (some papers reported several outcomes). Overall, 61 studies with 225799 males and 237017 females were eligible for meta-analysis. Among those, 20 papers with 141350 males and 165433 females fulfilled our criteria for odds ratio (OR) meta-analysis in seroprevalence, whereas the data from 41 papers (35 cohorts, 4 cross-sectional, and 2 case series) with 84449 males and 71584 females were used for mortality (66663 M, 52988 F), severity (19150 M, 19502 F) and recovery (43216 M, 32040 F) analyses. Tables 3 and 4 provide an overview of the included studies for meta-analysis.

**Table 3.**
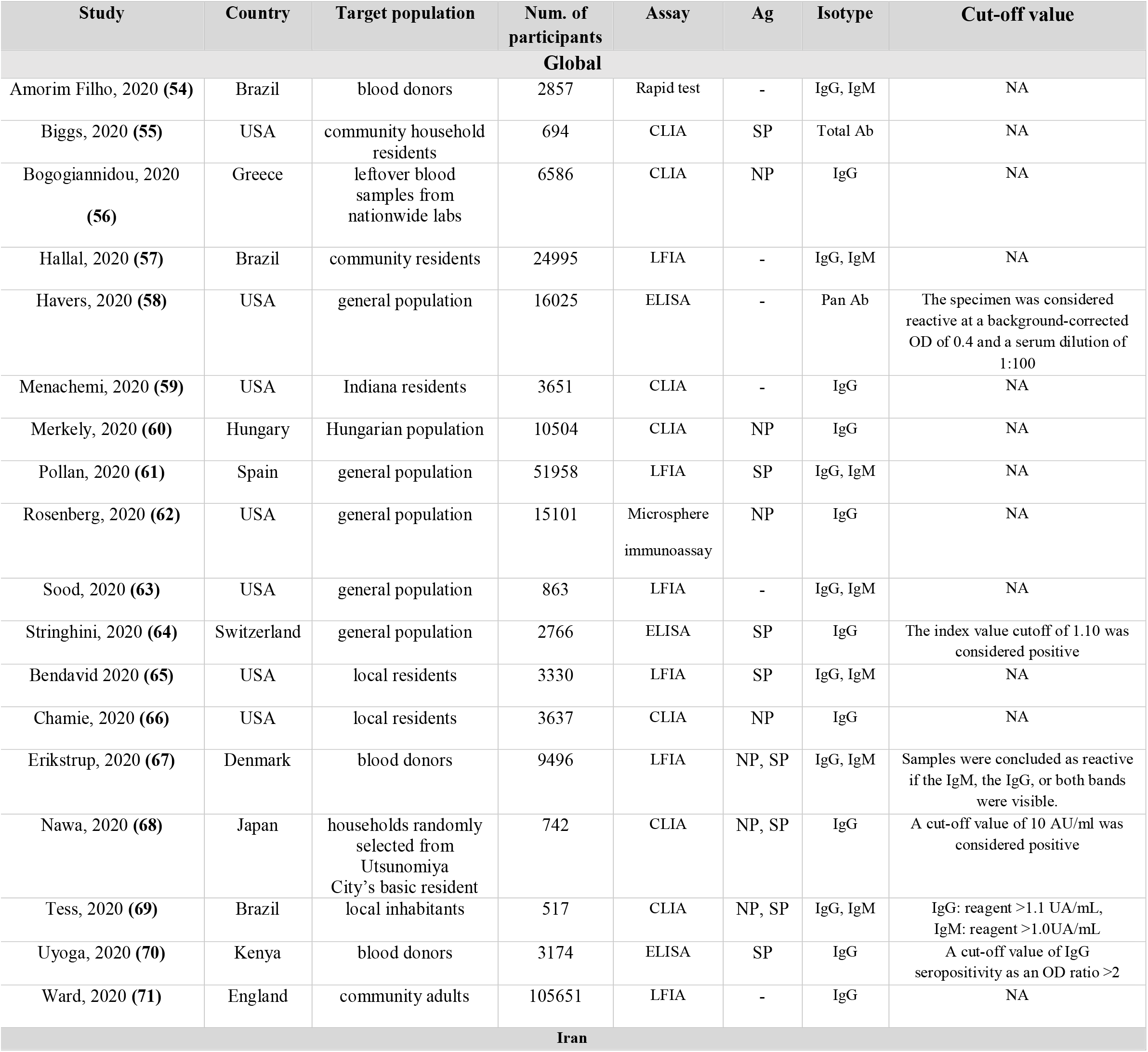
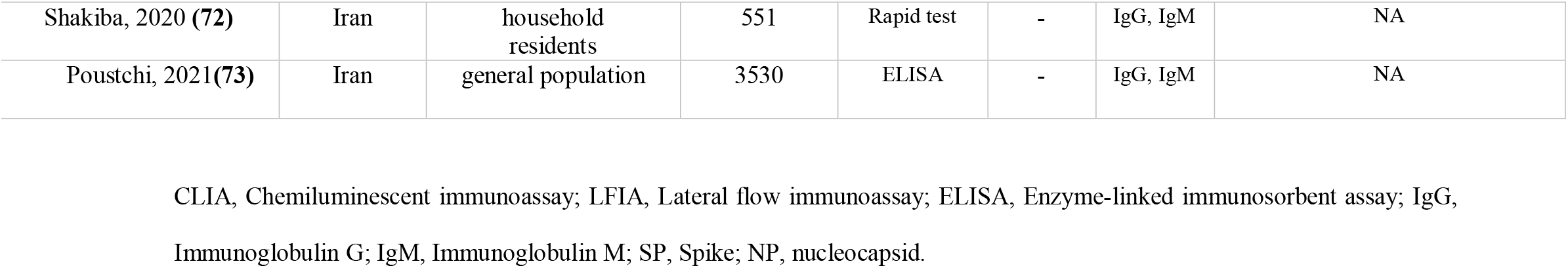
Included studies for seroprevalence meta-analysis.

**Table 4.**
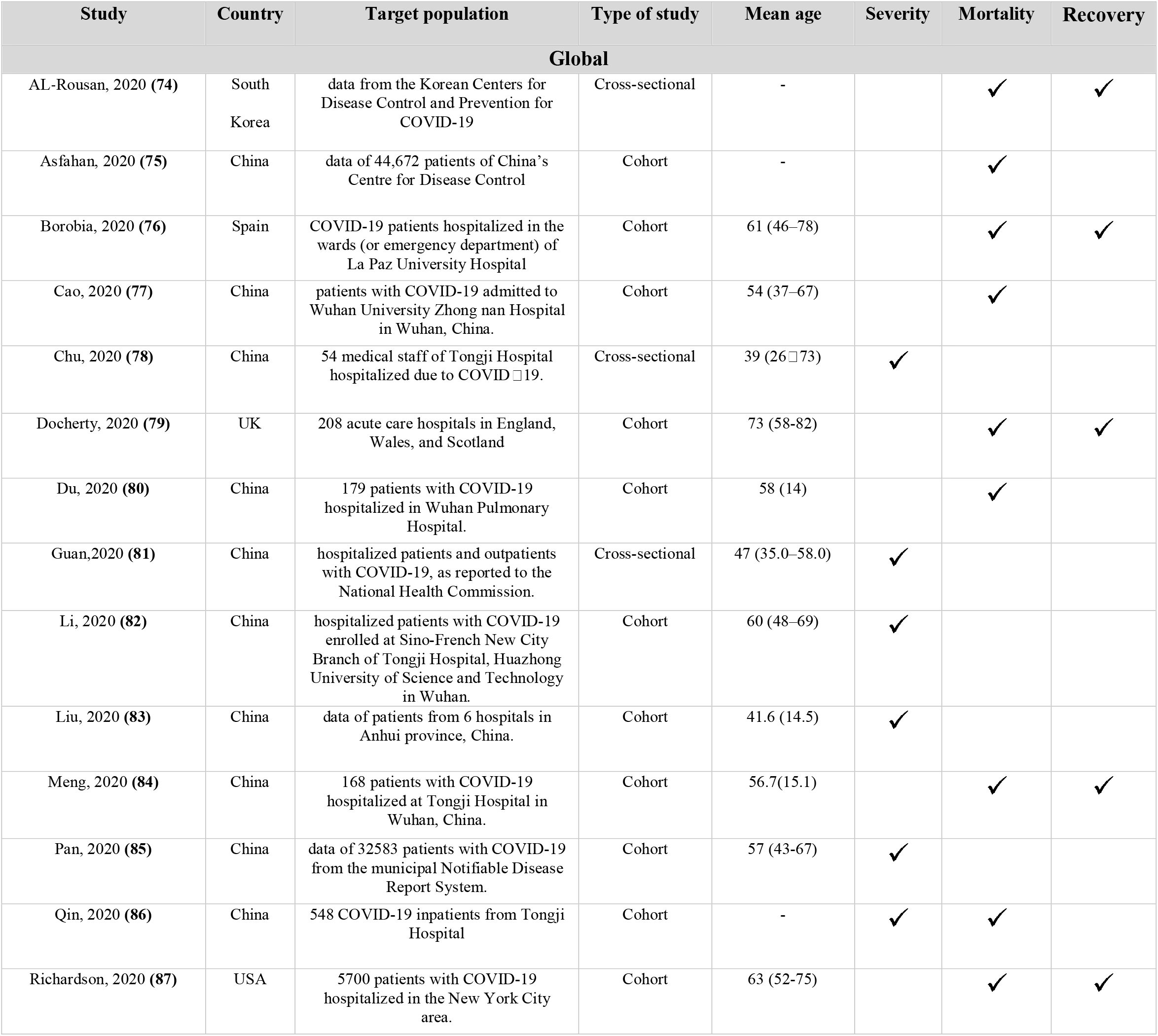
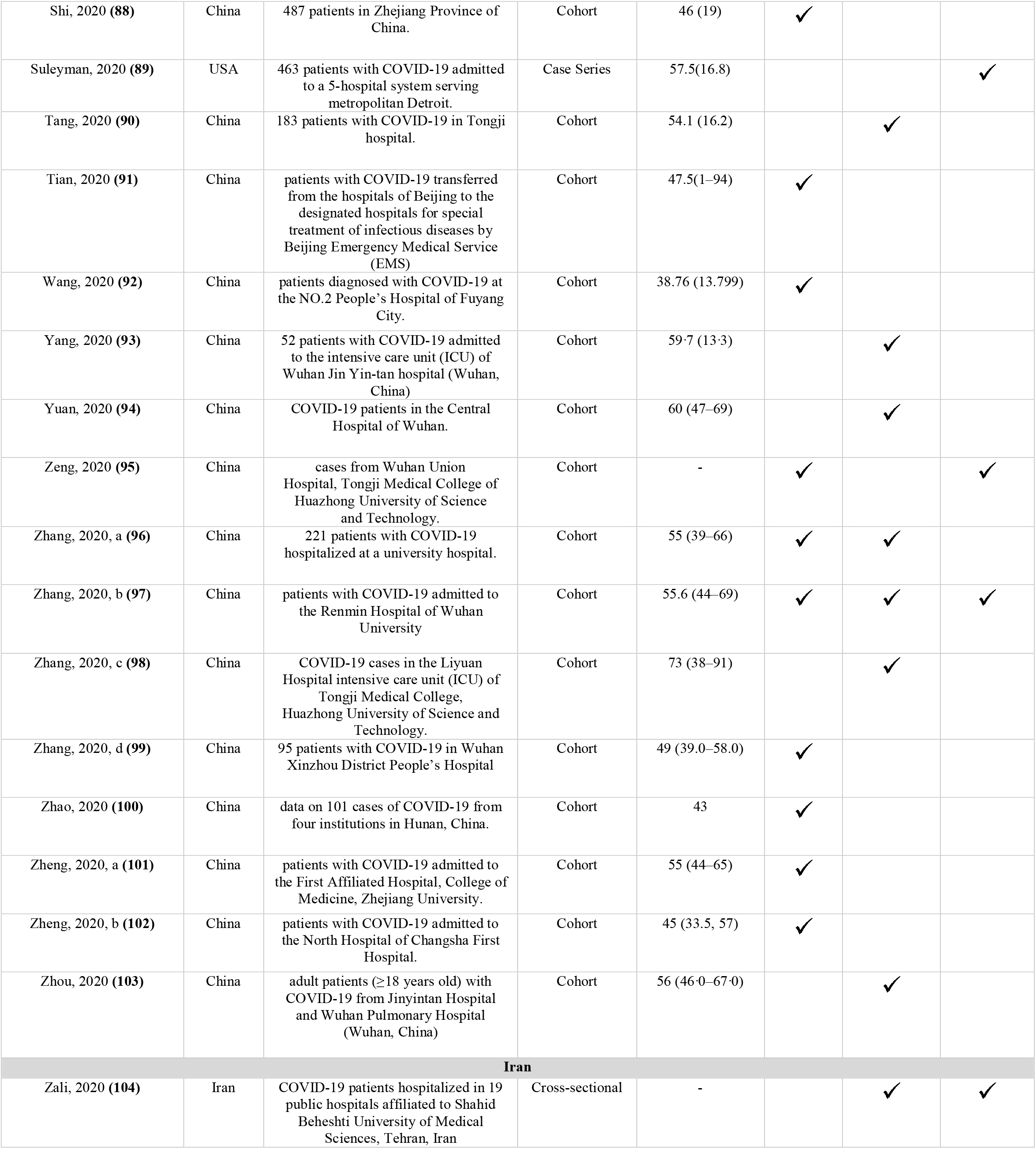
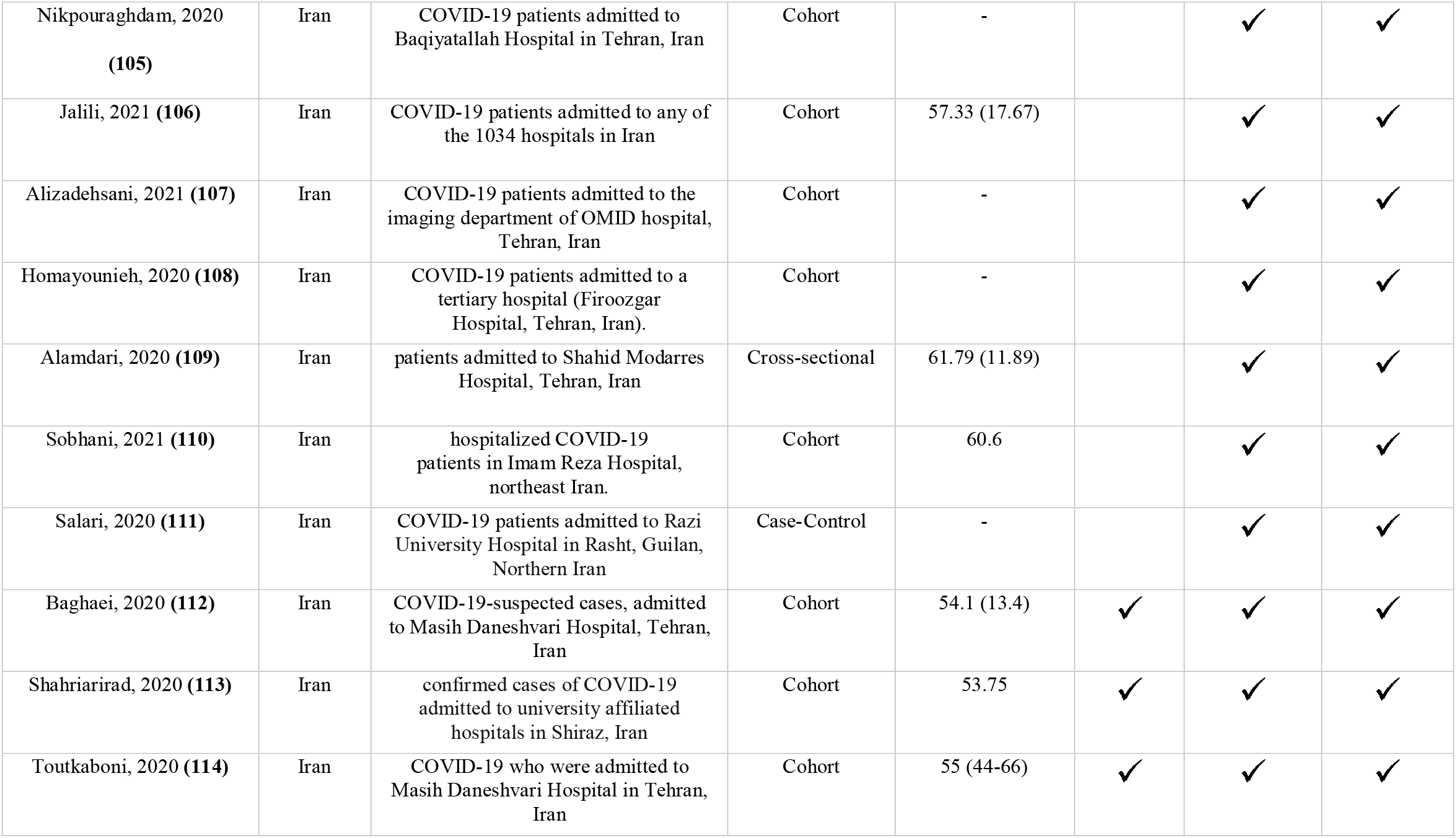
Included studies for severity, mortality, and recovery meta-analyses.

| Study | Country | Target population | Type of study | Mean age | Severity | Mortality | Recovery |
| --- | --- | --- | --- | --- | --- | --- | --- |
| **Global** |  |  |  |  |  |  |  |
| AL-Rousan, 2020 (74) | South Korea | data from the Korean Centers for Disease Control and Prevention for COVID-19 | Cross-sectional | - |  | ✓ | ✓ |
| Asfahan, 2020 (75) | China | data of 44,672 patients of China's Centre for Disease Control | Cohort | - |  | ✓ |  |
| Borobia, 2020 (76) | Spain | COVID-19 patients hospitalized in the wards (or emergency department) of La Paz University Hospital | Cohort | 61 (46–78) |  | ✓ | ✓ |
| Cao, 2020 (77) | China | patients with COVID-19 admitted to Wuhan University Zhong nan Hospital in Wuhan, China. | Cohort | 54 (37–67) |  | ✓ |  |
| Chu, 2020 (78) | China | 54 medical staff of Tongji Hospital hospitalized due to COVID-19. | Cross-sectional | 39 (26–73) | ✓ |  |  |
| Docherty, 2020 (79) | UK | 208 acute care hospitals in England, Wales, and Scotland | Cohort | 73 (58–82) |  | ✓ | ✓ |
| Du, 2020 (80) | China | 179 patients with COVID-19 hospitalized in Wuhan Pulmonary Hospital. | Cohort | 58 (14) |  | ✓ |  |
| Guan, 2020 (81) | China | hospitalized patients and outpatients with COVID-19, as reported to the National Health Commission. | Cross-sectional | 47 (35.0–58.0) | ✓ |  |  |
| Li, 2020 (82) | China | hospitalized patients with COVID-19 enrolled at Sino-French New City Branch of Tongji Hospital, Huazhong University of Science and Technology in Wuhan. | Cohort | 60 (48–69) | ✓ |  |  |
| Liu, 2020 (83) | China | data of patients from 6 hospitals in Anhui province, China. | Cohort | 41.6 (14.5) | ✓ |  |  |
| Meng, 2020 (84) | China | 168 patients with COVID-19 hospitalized at Tongji Hospital in Wuhan, China. | Cohort | 56.7(15.1) |  | ✓ | ✓ |
| Pan, 2020 (85) | China | data of 32583 patients with COVID-19 from the municipal Notifiable Disease Report System. | Cohort | 57 (43–67) | ✓ |  |  |
| Qin, 2020 (86) | China | 548 COVID-19 inpatients from Tongji Hospital | Cohort | - | ✓ | ✓ |  |
| Richardson, 2020 (87) | USA | 5700 patients with COVID-19 hospitalized in the New York City area. | Cohort | 63 (52–75) |  | ✓ | ✓ |
| Shi, 2020 (88) | China | 487 patients in Zhejiang Province of China. | Cohort | 46 (19) | ✓ |  |  |
| Suleyman, 2020 (89) | USA | 463 patients with COVID-19 admitted to a 5-hospital system serving metropolitan Detroit. | Case Series | 57.5(16.8) |  |  | ✓ |
| Tang, 2020 (90) | China | 183 patients with COVID-19 in Tongji hospital. | Cohort | 54.1 (16.2) |  | ✓ |  |
| Tian, 2020 (91) | China | patients with COVID-19 transferred from the hospitals of Beijing to the designated hospitals for special treatment of infectious diseases by Beijing Emergency Medical Service (EMS) | Cohort | 47.5(1–94) | ✓ |  |  |
| Wang, 2020 (92) | China | patients diagnosed with COVID-19 at the NO.2 People's Hospital of Fuyang City. | Cohort | 38.76 (13.799) | ✓ |  |  |
| Yang, 2020 (93) | China | 52 patients with COVID-19 admitted to the intensive care unit (ICU) of Wuhan Jin Yin-tan hospital (Wuhan, China) | Cohort | 59.7 (13.3) |  | ✓ |  |
| Yuan, 2020 (94) | China | COVID-19 patients in the Central Hospital of Wuhan. | Cohort | 60 (47–69) |  | ✓ |  |
| Zeng, 2020 (95) | China | cases from Wuhan Union Hospital, Tongji Medical College of Huazhong University of Science and Technology. | Cohort | - | ✓ |  | ✓ |
| Zhang, 2020, a (96) | China | 221 patients with COVID-19 hospitalized at a university hospital. | Cohort | 55 (39–66) | ✓ | ✓ |  |
| Zhang, 2020, b (97) | China | patients with COVID-19 admitted to the Renmin Hospital of Wuhan University | Cohort | 55.6 (44–69) | ✓ | ✓ | ✓ |
| Zhang, 2020, c (98) | China | COVID-19 cases in the Liyuan Hospital intensive care unit (ICU) of Tongji Medical College, Huazhong University of Science and Technology. | Cohort | 73 (38–91) |  | ✓ |  |
| Zhang, 2020, d (99) | China | 95 patients with COVID-19 in Wuhan Xinzhou District People's Hospital | Cohort | 49 (39.0–58.0) | ✓ |  |  |
| Zhao, 2020 (100) | China | data on 101 cases of COVID-19 from four institutions in Hunan, China. | Cohort | 43 | ✓ |  |  |
| Zheng, 2020, a (101) | China | patients with COVID-19 admitted to the First Affiliated Hospital, College of Medicine, Zhejiang University. | Cohort | 55 (44–65) | ✓ |  |  |
| Zheng, 2020, b (102) | China | patients with COVID-19 admitted to the North Hospital of Changsha First Hospital. | Cohort | 45 (33.5, 57) | ✓ |  |  |
| Zhou, 2020 (103) | China | adult patients (≥18 years old) with COVID-19 from Jinyintan Hospital and Wuhan Pulmonary Hospital (Wuhan, China) | Cohort | 56 (46.0–67.0) |  | ✓ |  |
| **Iran** |  |  |  |  |  |  |  |
| Zali, 2020 (104) | Iran | COVID-19 patients hospitalized in 19 public hospitals affiliated to Shahid Beheshti University of Medical Sciences, Tehran, Iran | Cross-sectional | - |  | ✓ | ✓ |
| Nikpouraghdam, 2020<br>**(105)** | Iran | COVID-19 patients admitted to Baqiyatallah Hospital in Tehran, Iran | Cohort | - |  | ✓ | ✓ |
| Jalili, 2021 **(106)** | Iran | COVID-19 patients admitted to any of the 1034 hospitals in Iran | Cohort | 57.33 (17.67) |  | ✓ | ✓ |
| Alizadehsani, 2021 **(107)** | Iran | COVID-19 patients admitted to the imaging department of OMID hospital, Tehran, Iran | Cohort | - |  | ✓ | ✓ |
| Homayounieh, 2020 **(108)** | Iran | COVID-19 patients admitted to a tertiary hospital (Firoozgar Hospital, Tehran, Iran). | Cohort | - |  | ✓ | ✓ |
| Alamdari, 2020 **(109)** | Iran | patients admitted to Shahid Modarres Hospital, Tehran, Iran | Cross-sectional | 61.79 (11.89) |  | ✓ | ✓ |
| Sobhani, 2021 **(110)** | Iran | hospitalized COVID-19 patients in Imam Reza Hospital, northeast Iran. | Cohort | 60.6 |  | ✓ | ✓ |
| Salari, 2020 **(111)** | Iran | COVID-19 patients admitted to Razi University Hospital in Rasht, Guilan, Northern Iran | Case-Control | - |  | ✓ | ✓ |
| Baghaei, 2020 **(112)** | Iran | COVID-19-suspected cases, admitted to Masih Daneshvari Hospital, Tehran, Iran | Cohort | 54.1 (13.4) | ✓ | ✓ | ✓ |
| Shahriarirad, 2020 **(113)** | Iran | confirmed cases of COVID-19 admitted to university affiliated hospitals in Shiraz, Iran | Cohort | 53.75 | ✓ | ✓ | ✓ |
| Toutkaboni, 2020 **(114)** | Iran | COVID-19 who were admitted to Masih Daneshvari Hospital in Tehran, Iran | Cohort | 55 (44-66) | ✓ | ✓ | ✓ |

**Figure 1.**
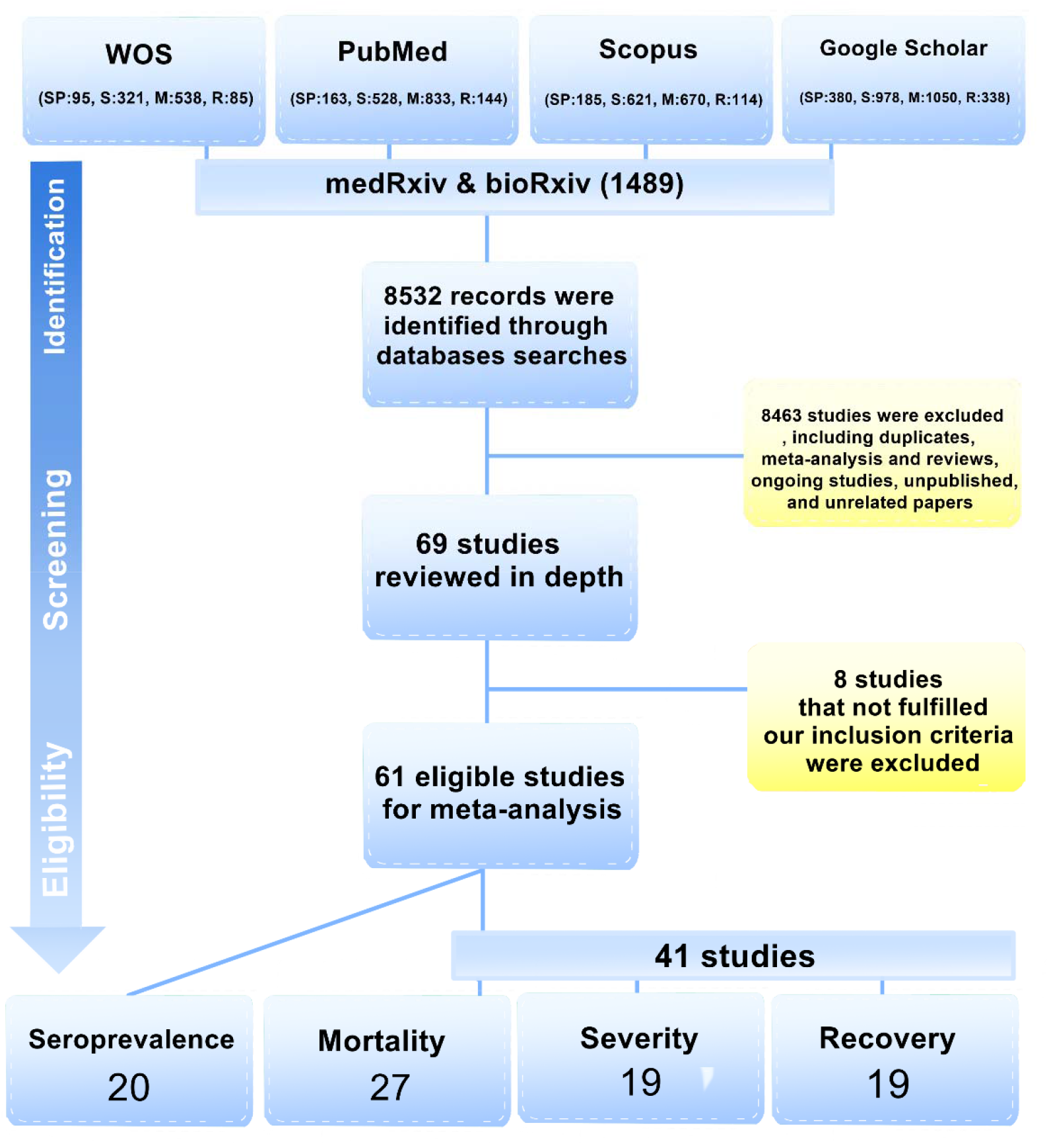
PRISMA flow diagram of the study selection process.

### 3.2 Meta-analysis of seroprevalence

Nine out of twenty seroprevalence studies measured only IgG, nine reported both IgG and IgM, and two studies measured total antibodies. Multiple detection methods were used in the studies, including chemiluminescent immunoassay (CLIA), lateral flow immunoassay (LFIA), and enzyme-linked immune sorbent assay (ELISA). The largest number of included papers for seroprevalence analyses was from the USA (seven studies), whereas the largest number of included papers for other analyses was from China. Comparing males and females, who were RT-PCR negative for COVID-19, showed that there was a significant difference in the global seroprevalence rate. Analysis indicated that males (4395 out of 139285) were 13% more seropositive than females (5071 out of 163417) in the general population [OR 1.13 (95% CI: 1.03, 1.24), *p*-value =0.009, Fig. 2a]. However, no significant difference was seen in the seroprevalence rate between males and females in Iran [OR 0.93 (95% CI: 0.79, 1.11), *p*-value =0.43, Fig. 2b]. For exploring differences in seroprevalence based on age, subgroup analyses were performed for three subgroups [young (Y), <40 years; middle-aged (M), 40-60 years; old (O), >60 years]. The results showed no significant difference between young, middle-aged, and old individuals in terms of seroprevalence rate [(OR for Y/M=0.88, 95% CI: 0.74 to 1.04, *p=* 0.14, Fig 3a) (OR for O/M= 0.85, 95% CI: 0.61 to 1.18, *p=* 0.32, Fig 3b) (OR for Y/O= 1.00, 95% CI: 0.71 to 1.42, *p=* 0.98, Fig 3c)]. Subgroup analyses according to the type of antibody and detection assay were also performed. Antibody isotype-based analyses confirmed the higher seroprevalence rate in men, but the differences were not significant for both IgG and IgG/IgM, ([OR]=1.13, 95% [CI]: 0.98 to 1.31, *p=* 0.09, Fig 4a) and ([OR] = 1.05, 95% [CI]: 0.95 to 1.16, *p=* 0.33, Fig 4b), respectively. Total subgroups analyses based on the detection method showed similar results for CLIA, LFIA, and ELISA assays in males and females, ([OR]=1.11, 95% [CI]: 0.88 to 1.41, *p=* 0.38, Fig 4c) and ([OR] = 1.01, 95% [CI]: 0.94 to 1.08, *p=* 0.78, Fig 4d) and ([OR]=1.13, 95% [CI]: 0.93 to 1.37, *p=* 0.23, Fig 4e).

**Figure 2.**
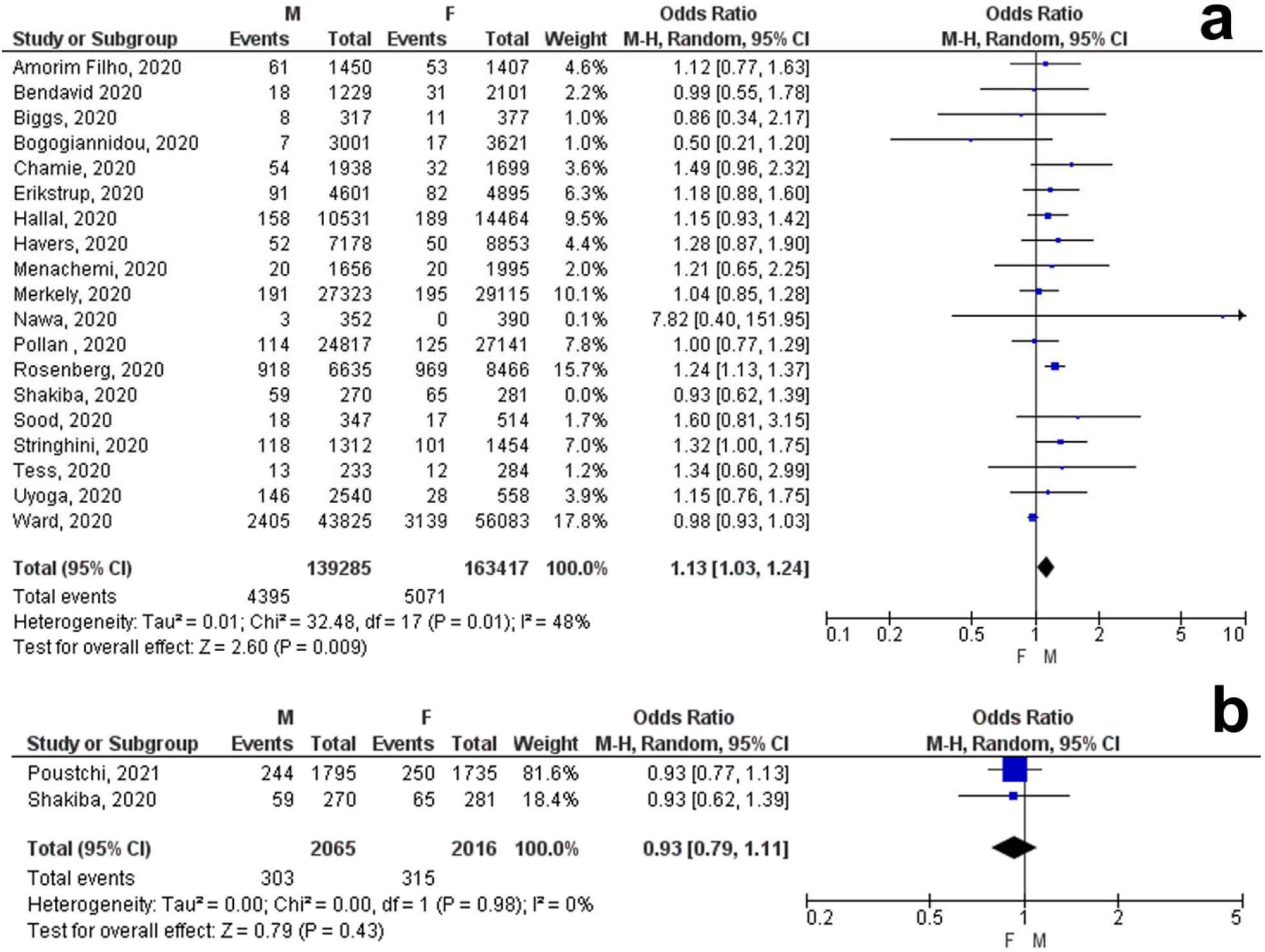
Gender difference meta-analysis of COVID-19 seroprevalence in included studies. Global (a), Iran (b).

**Figure 3.**
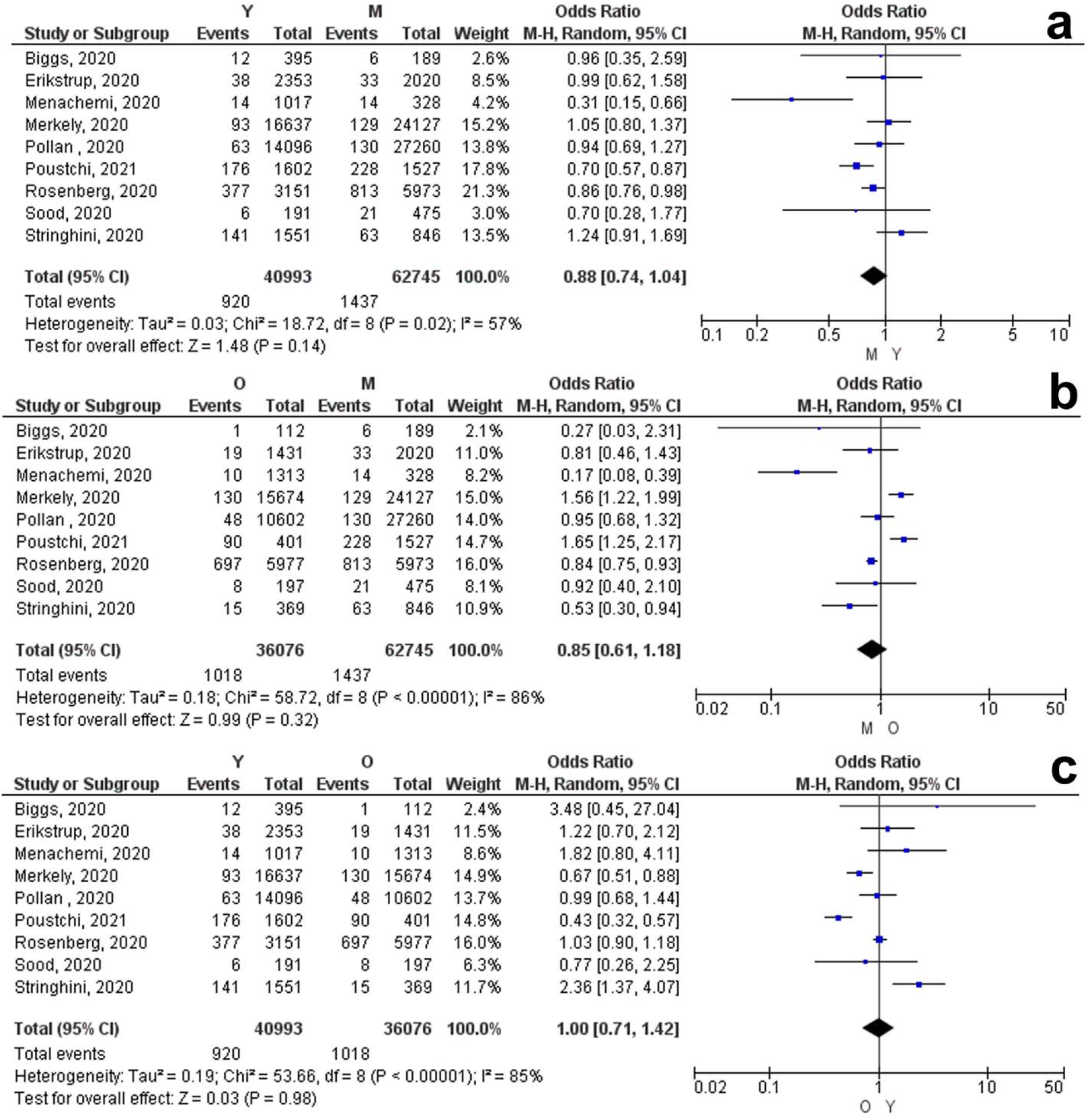
Subgroup meta-analysis based on age for COVID-19 seroprevalence. Young vs. middle-aged (a), elderly vs. middle-aged (b), young vs. elderly (c).

**Figure 4.**
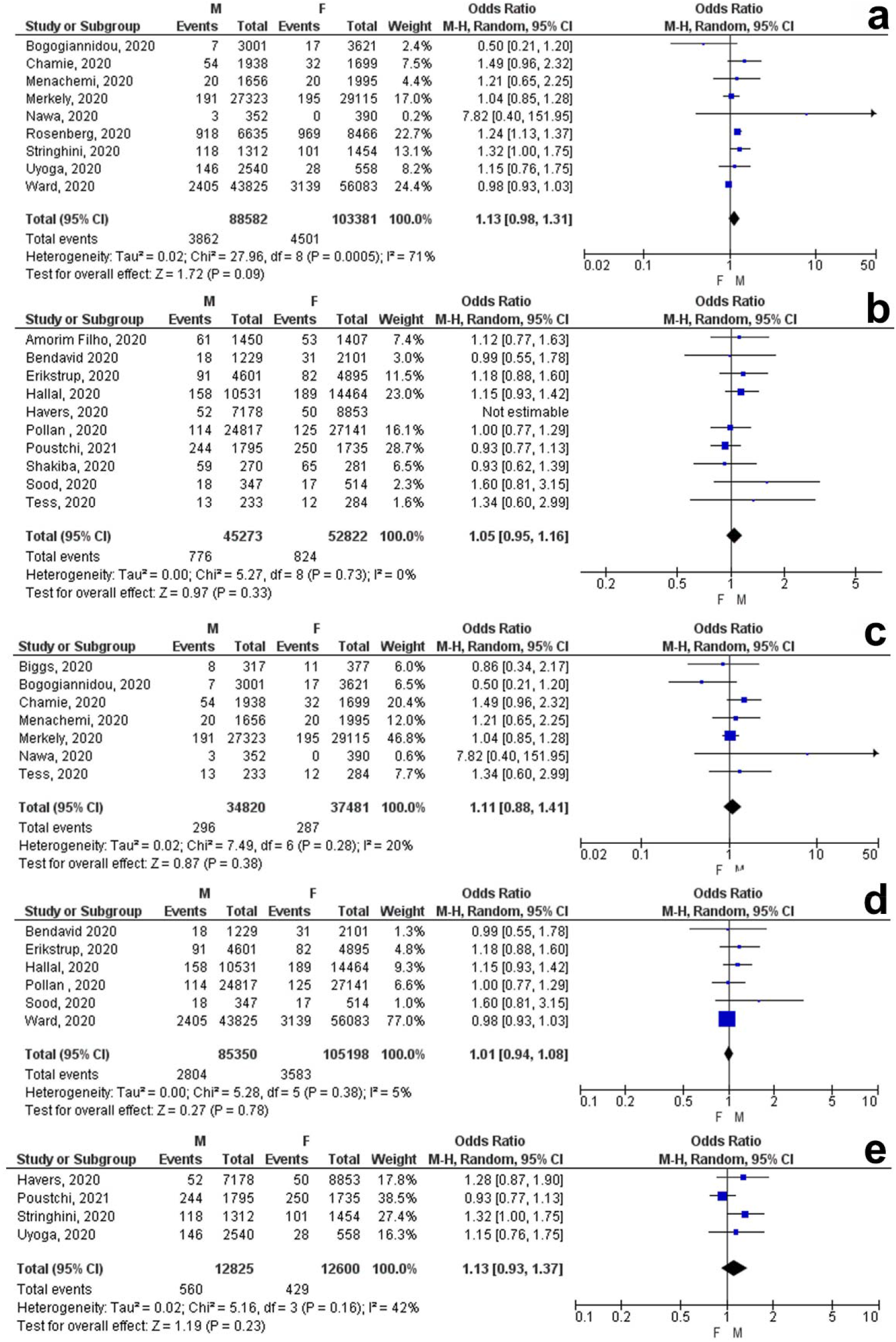
Subgroup meta-analysis based on antibody isotype and detection assay for COVID-19 seroprevalence. IgG (a), IgG/IgM (b), CLIA (c), LFIA (d), ELISA (e).

### 3.3 Meta-analysis of mortality

Sex-specific comparison in COVID-19 confirmed cases across the globe (76844 patients) indicated that there was a significant difference in mortality between males and females, which was 45% higher in men (4789 out of 41982) than women (2792 out of 34862 individuals) [OR 1.45 (95% CI: 1.19, 1.77), *p*-value <0.001, Fig. 5a]. The results of the meta-analysis for mortality of Iranian patients were in line with the global pattern (4612 death in 24681 M and 2858 death in 18126 F) and 26% higher in men [OR 1.26 (95% CI: 1.20, 1.33), *p*-value <0.001, Fig. 5b]. For exploring associations between mortality and age, subgroup analysis was performed based on mean (SD) comparison between victims and survivors. The results demonstrated a significant standardized mean difference of death by 2.71 years [95% [CI]: 1.44 to 3.98, *p<* 0.001, Fig 5c), while this difference was just 0.73 years for Iranian patients [95% [CI]: 0.54 to 0.92, *p<* 0.001, Fig 5d).

**Figure 5.**
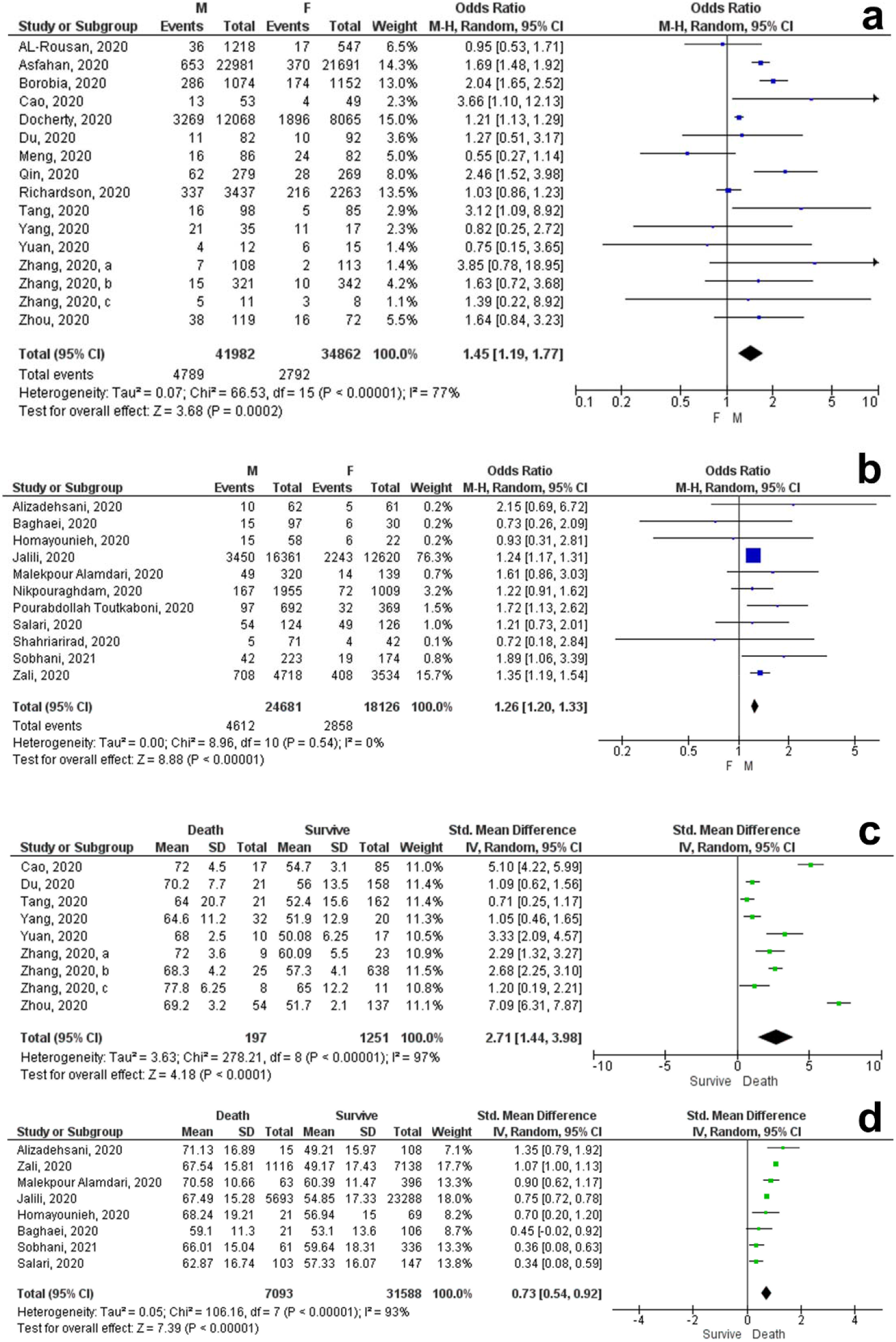
Gender difference meta-analysis of COVID-19 mortality in included studies. Mortality based on sex: Global (a), Iran (b). mortality based on mean age (SD): Global (c), Iran (d).

**Figure 6.**
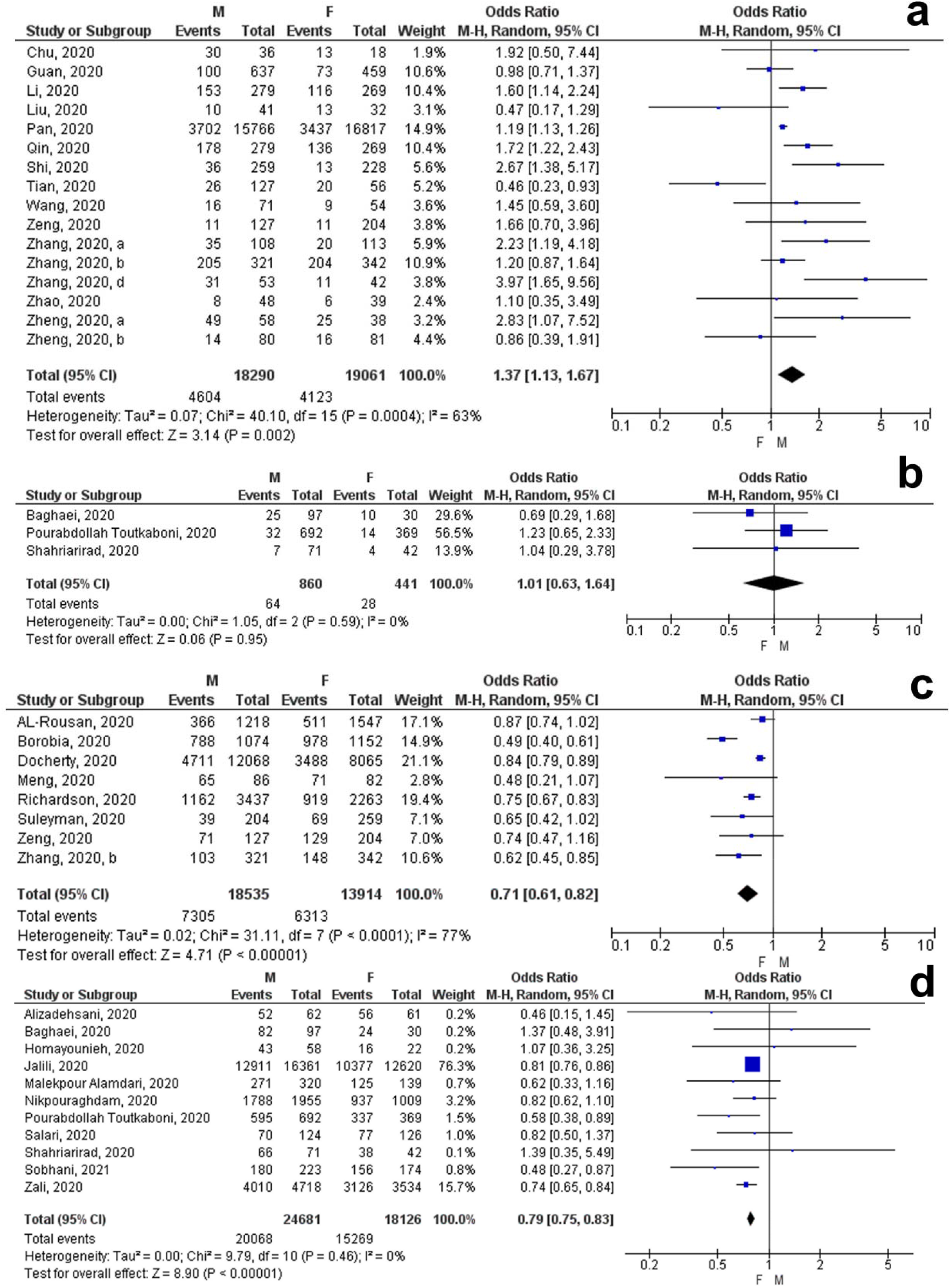
Gender difference meta-analysis of COVID-19 severity and recovery in included studies. Severity: Global (a), Iran (b). Recovery and discharge: Global (c), Iran (d).

### 3.4 Meta-analysis of severity and recovery

Sex-based meta-analysis in COVID-19 confirmed cases (37351 patients) showed that the disease is 37% more severe in men (4604 out of 18290) than women in the global pattern (4123 out of 19061) [OR 1.37 (95% CI: 1.13, 1.67), *p*-value =0.002, Fig. 5a]. Although, there was no significant difference in the severity of the disease between Iranian men and women [OR 1.01 (95% CI: 0.63, 1.64), *p*-value =0.95, Fig. 5b]. As expected, the rate of recovery and hospital discharge in males (7305 out of 18535) were 29% lower than females in the global pattern (6313 out of 13914 individuals) [OR 0.71 (95% CI: 0.61, 0.82), *p*-value <0.001, Fig. 5c]. The recovery rate is also 21% higher in Iranian women (15269 out of 18126 individuals) compared to men (20068 out of 24681 individuals) [OR 0.79 (95% CI: 0.54, 0.92), *p*-value <0.001, Fig. 5d].

### 3.6 Risk of bias and quality assessment

Studies’ risk of biases is provided in Table 5-6. The Newcastle-Ottawa Scale was used to assess the overall quality. For the included studies to evaluate seroprevalence, three criteria were evaluated, including the study population and participants’ characteristics, detection assay, and outcomes. For the rest of the studies the type of study and study population, the comparability, and the outcomes were evaluated. Most of the included studies had moderate to high quality of evidence. Besides, a total of twelve studies were evaluated as low quality.

**Table 5.** Risk of bias of included studies for seroprevalence.

| Study | Newcastle-Ottawa Score |  |  |  |
| --- | --- | --- | --- | --- |
|  | Study population and characteristics (max=3) | Detection method and validation (max=3) | Outcomes (max=4) | Total quality (L<6, M=7-8, H>8) |
| Amorim Filho, 2020 | 1 | 1 | 3 | Low |
| Biggs, 2020 | 3 | 3 | 2 | Moderate |
| Bogogiannidou, 2020 | 0 | 3 | 4 | Moderate |
| Hallal, 2020 | 3 | 3 | 3 | High |
| Havers, 2020 | 1 | 2 | 4 | Moderate |
| Menachemi, 2020 | 3 | 0 | 2 | Low |
| Merkely, 2020 | 3 | 1 | 2 | Low |
| Pollan, 2020 | 3 | 3 | 2 | Moderate |
| Poustchi, 2021 | 3 | 3 | 3 | High |
| Rosenberg, 2020 | 1 | 3 | 4 | Moderate |
| Sood, 2020 | 3 | 3 | 4 | High |
| Stringhini, 2020 | 3 | 2 | 4 | High |
| Bendavid 2020 | 1 | 3 | 4 | Moderate |
| Chamie, 2020 | 1 | 1 | 4 | Low |
| Erikstrup, 2020 | 1 | 3 | 2 | Low |
| Nawa, 2020 | 2 | 1 | 2 | Low |
| Shakiba, 2020 | 3 | 1 | 4 | Moderate |
| Tess, 2020 | 2 | 3 | 2 | Moderate |
| Uyoga, 2020 | 1 | 2 | 4 | Moderate |
| Ward, 2020 | 2 | 1 | 4 | Moderate |

**Table 6.** Risk of bias of included studies for severity, mortality, and recovery.

| Study | Newcastle-Ottawa score |  |  |  |
| --- | --- | --- | --- | --- |
|  | Study population and type of study (max=4) | Comparability (max=3) | Outcomes (max=3) | Total quality (L<6, M=7-8, H>8) |
| AL Rousan, 2020 | 2 | 2 | 2 | Low |
| Alamdari, 2020 | 2 | 2 | 2 | Low |
| Alizadehsani, 2021 | 3 | 2 | 2 | Moderate |
| Asfahan, 2020 | 3 | 2 | 2 | Moderate |
| Baghaei, 2020 | 3 | 1 | 2 | Low |
| Borobia, 2020 | 3 | 3 | 2 | Moderate |
| Cao, 2020 | 3 | 1 | 3 | Moderate |
| Chu, 2020 | 2 | 1 | 2 | Low |
| Docherty, 2020 | 3 | 2 | 2 | Moderate |
| Du, 2020 | 4 | 2 | 3 | High |
| Guan, 2020 | 2 | 1 | 3 | Low |
| Homayounieh, 2020 | 3 | 2 | 2 | Moderate |
| Jalili, 2021 | 3 | 3 | 3 | High |
| Li, 2020 | 4 | 3 | 3 | High |
| Liu, 2020 | 3 | 1 | 2 | Low |
| Meng, 2020 | 3 | 3 | 3 | High |
| Nikpouraghdam, 2020 | 3 | 2 | 2 | Moderate |
| Pan, 2020 | 4 | 3 | 3 | High |
| Qin, 2020 | 3 | 2 | 3 | Moderate |
| Richardson, 2020 | 4 | 1 | 3 | Moderate |
| Salari, 2020 | 2 | 1 | 1 | Low |
| Shi, 2020 | 3 | 1 | 1 | Low |
| Shahriarirad, 2020 | 3 | 2 | 2 | Moderate |
| Sobhani, 2021 | 3 | 2 | 2 | Moderate |
| Suleyman, 2020 | 2 | 2 | 2 | Low |
| Tang, 2020 | 4 | 1 | 3 | Moderate |
| Tian, 2020 | 4 | 1 | 2 | Moderate |
| Toutkaboni, 2020 | 3 | 1 | 1 | Low |
| Wang, 2020 | 3 | 2 | 2 | Moderate |
| Yang, 2020 | 4 | 3 | 2 | High |
| Yuan, 2020 | 4 | 1 | 3 | Moderate |
| Zali, 2020 | 2 | 2 | 2 | Low |
| Zeng, 2020 | 4 | 1 | 2 | Moderate |
| Zhang, 2020, a | 4 | 1 | 3 | Moderate |
| Zhang, 2020, b | 4 | 3 | 3 | High |
| Zhang, 2020, c | 4 | 1 | 2 | Moderate |
| Zhang, 2020, d | 3 | 1 | 3 | Moderate |
| Zhao, 2020 | 3 | 1 | 2 | Moderate |
| Zheng, 2020, a | 4 | 1 | 3 | Moderate |
| Zheng, 2020, b | 4 | 1 | 2 | Moderate |
| Zhou, 2020 | 4 | 1 | 2 | Moderate |

## 4. DISCUSSION

Since the introduction of serological diagnostic kits for SARS-CoV-2 and their availability to researchers around the world, various studies and papers have been published to investigate seroprevalence rates among various ethnic populations (13). These types of studies can be a very valuable screenshot to check the immune status as well as the previous exposure to the virus. However, many factors can affect seroprevalence in the general population, including sampling time, epidemic status in the studied country, use of personal protective equipment (PPE), type of targeted antigen, type of detection assay, antibody isotype, and its cut-off value. Although the WHO has defined a population-based serological study protocol (14), our searches showed that, to date, at least a small number of studies have met these standard protocols (13). For this reason, heterogeneity could be high in our reviewed published papers. In addition to the subgroup analyses which were performed in the present study, every effort was made to include studies that have the maximum similarity for meta-analyses. In order to reduce the impact of jobs, exposure, and the use of PPE on main outcomes, only studies conducted on the general population were reviewed. However, the information reported in the papers on how and to what extent possible exposure to the virus was very incomplete, and this was one of the major limitations we faced.

The results of our pooled analysis showed that there was a significant difference in seroprevalence rate between males and females which males 13% more likely to be seropositive than females in the general population. Although the results of the meta-analysis were not significant for seroprevalence among the Iranian population, it did not seem to follow the global pattern and the prevalence of anti-COVID-19 Ab was higher in women (7%). Of course, this difference can be due to the variations in the number of articles included for the analysis. There was no significant difference in subgroup analyses based on age, antibody isotype, and detection assays. The difference between male and female seroprevalence in the global pattern can be due to several reasons. First, males may be more exposed to COVID-19 than women and may use less PPE (15, 16). Second, based on the previous knowledge about stronger immune responses in females, perhaps the reason for the lower seroprevalence in them is the stronger innate immune response including type 1 interferon at the onset of the disease (17-19). Although, the results obtained from analyses in Iranian population showed that this pattern is not similar in all countries and regions. Besides, no significant difference was observed in seroprevalence analyses based on age, which is an effective factor in the rate of exposure. A recent systematic review study reported a positive association between seroprevalence and age among participants younger than 65 years and no significant difference between males and females (20), unlike our study. However, this study was performed on all populations, including healthcare workers and close contacts, and was not limited to the general population.

Consistent with the results of two recent systematic reviews (21, 22), we have also found that males have more severe disease and a worse prognosis than females and their mortality rate is up to 45% higher than females. The results of the analysis on the Iranian population also showed a similar pattern, albeit with a milder slope (26%). The meta-analyses also showed that the age difference between the dead and rescued people in the Iranian population is only 0.73 years, while this number is 2.71 years for other countries.

Our present study also examined the number of men and women who were recovered and discharged from the hospital, and the results showed that, as expected, males have up to 29% less recovery and discharge than females. The results in the Iranian population, in line with the global pattern, also showed a recovery of up to 21% more in women than men. There are several reasons for this situation that have not been proven with certainty to date. Studies have shown that Angiotensin-converting enzyme 2 (ACE2), expressed on the PAR region of the X chromosome, is regulated by estrogen (23). *Lambert et al*. showed in their study on SARS-CoV-1 that females may express more circulating ACE2 which can protect them against acute respiratory distress syndrome (24). Besides, both humoral and cellular immunity is stronger in women (25), and it has been shown that early IFN responses and the production of neutralizing antibodies are highly effective in reducing disease severity (8, 26). But as shown in Tables 1 and 2, there are conflicting results about the effects of estrogen as the main female hormone on ACE2 expression. Also, despite the proven strong antiviral effects of interferons, especially interferon type 1, it is still not possible to comment with certainty on its role in gender differences.

## 5. CONCLUSION

In summary, by aggregating the results of meta-analyses for seroprevalence, severity, mortality, and recovery, it is clear that there are gender differences in COVID-19. Also, comparing the mortality and recovery rate in global pattern with Iran showed similar results. However, there seem to be interesting differences in serum prevalence between different populations of the world. The reason for this difference is not completely understood, but what is clear and has been proven in many autoimmune diseases is that there are sex biases in immune responses. Therefore, it seemed that a new approach should be taken in the fight against COVID-19 and the population of men, especially older men, should be further studied and cared for. Besides, the results of this study and similar studies can be a warning to those who have started a dangerous game with herd immunity regardless of scientific support. Hoping for COVID-19-free days.

## Data Availability

The authors confirm that the data supporting the findings of this study are available within the manuscript.

## 6. ABBREVIATIONS

ACE2: Angiotensin-converting enzyme 2
CLIA: Chemiluminescent immunoassay
CI: Confidence interval
COVID-19: Coronavirus disease 2019
ELISA: Enzyme-linked immunosorbent assay
IgG: Immunoglobulin G
IgM: Immunoglobulin M
LFIA: Lateral flow immunoassay
OR: odds ratio
PPE: Personal protective equipment
RT-PCR: Real-time polymerase chain reaction
SARS-CoV-2: Severe acute respiratory syndrome coronavirus 2
SD: Standard deviation
WHO: World Health Organization

## 7. DECLARATIONS

### 7.1 Ethical Approval and Consent to participate

Not applicable.

### 7.2 Consent for publication

Not applicable.

### 7.3 Availability of supporting data

Not applicable.

### 7.6 Authors’ contributions

HAO and MRN conceived the study and wrote the manuscript. MRN performed the literature search and selected the relevant studies. HAO and MRN provided methodological support in the study design and classified retrieved articles according to the level of evidence. HAO critically revised the manuscript and provided the final approval.

## 7.7 Acknowledgements

The authors thank the immunology department of Mazandaran University of Medical Sciences for their kind and prompt support.

